# The influence of Quality-of-Life Orientation on Recovery College advertising: A Dutch-English-Japanese corpus-based comparison

**DOI:** 10.1101/2025.08.13.25333552

**Authors:** Yasuhiro Kotera, Marijn Antens, Cornelis L. Mulder, Stynke Castelein, Yuki Miyamoto, Ruth Rietveld, Agnieszka Kapka, Amy Ronaldson, Simran Takhi, Tesnime Jebara, Michio Murakami, Dan Elton, Peter Bates, Claire Henderson, Mike Slade, Sara Vilar-Lluch

## Abstract

**Background:** Recovery Colleges (RCs) aim to democratise mental health support through co-produced education. While increasingly adopted worldwide, their public-facing communications may reflect culturally specific values.

**Aim:** This study examined (a) the textual emphases in how RCs are presented to the public in the Netherlands, and (b) how the cultural dimension of Quality-of-Life Orientation is reflected in these texts, through comparison with those from England and Japan.

**Methods:** Promotional texts (3,445 words) from 25 Dutch RCs were analysed using corpus-based discourse analysis (Wordlist, Word Sketch, and Keywords) and compared to existing corpora from England (61 RCs; 22,014 words) and Japan (13 RCs; 813 words). Interpretations were informed by Hofstede’s Cultural Dimensions Theory.

**Results:** Dutch RC texts foregrounded recovery as a personal yet socially contextualised process, emphasising agency, vulnerability, and participation in society. Unlike English RCs, educational terminology was largely absent, and the concept of “academy” or “meeting place” was prominent. Compared to the individualised framing in England and the collectivist emphasis in Japan, Dutch RCs represented a midpoint, highlighting recovery in everyday life and reflecting a culturally embedded Quality-of-Life Orientation.

**Conclusion:** These findings suggest that cultural orientations shape the language of RC advertising, which may influence public expectations and, in turn, how RCs are understood and operated. Such dynamics are important to consider in international RC development and cultural adaptation, to ensure culturally resonant communication and engagement.

## Introduction

Mental health personal recovery is commonly regarded as a deeply personal and unique process involving changes in attitudes, values, feelings, goals, and roles, enabling individuals to live fulfilling lives despite ongoing mental health challenges (1). It has gained considerable traction within mental health systems worldwide (2). Recovery-oriented approaches have been adopted globally and shown to promote hope, empowerment, autonomy, and social inclusion among service users, while also improving attitudes and understanding among mental health professionals (3). As a result, recovery principles have been embedded into policy frameworks across various national policies (4–11).

Recovery Colleges (RCs) represent a novel recovery-oriented approach that blends social support with co-produced, adult-learning programmes for service users, carers, and professionals. Originating in England in 2009 and influenced by earlier U.S. peer-run services, RCs have since expanded to 28 countries across diverse cultural and economic settings (2, 12). RCs operate across various service contexts, such as health services, NGOs, and educational institutions, and are recognised as a mental health innovation (13). Central to their model are co-production and adult education: co-production involves lived experience and professional expertise in all stages of course delivery (14), while adult education promotes inclusive, strengths-based, and self-directed learning (15).

Together, these approaches aim to foster personal recovery, defined by living a meaningful and autonomous life despite symptoms (2). In the RC model, this is facilitated through social inclusion and the development of life skills. RCs are open to anyone, including people with lived experience and their carers, and courses cover a range of topic such as self-management of wellbeing, mental health conditions and symptoms, creativity, physical health, social connection and practical life skills (16).

Evidence supporting the effectiveness and cost-effectiveness of RCs is growing. Reviews and qualitative syntheses have highlighted benefits for students (e.g., improved confidence, hope, empowerment, and reduced stigma) and for staff (e.g., increased motivation and positive attitudinal shifts) (15, 17, 18). Economically, RC participation has been linked to reduced service use and net savings, such as AU$269 per student annually in Australia (19, 20).

However, cross-cultural evaluation of RCs remains underdeveloped (21). To advance cross-cultural understanding of RCs, global studies have recently been conducted, involving the 28 RC-operating countries (22, 23). These studies found that the current RC operational model, informed by key RC fidelity components (24), aligns more closely with the cultural characteristics of WEIRD countries, based on Hofstede’s Cultural Dimensions Theory (25). Table 1 explains all six dimensions. Of the six dimensions, four—Individualism, Indulgence, Uncertainty Acceptance, and Short-Term Orientation—were significantly associated with the RC operational model; that is, RCs in countries characterised by these cultural dimensions tended to score higher on RC operation fidelity (23). These findings indicate the importance of cross-cultural investigation in RCs.

**Table 1.** Six cultural dimensions in the Cultural Dimension Theory.

| **Characteristic (Interpretation)** | **Meaning** |
| --- | --- |
| Individualism* (vs Collectivism) | A degree to which a society expects individuals to be loosely tied to one another, and to take care of only themselves and their immediate family. |
| Indulgence* (vs Self-Restrained) | Acceptance of relatively free gratification of basic and natural human needs to enjoy life. |
| Uncertainty Avoidance (vs Uncertainty Acceptance*) | A degree to which individuals feel threatened by unknown situations, and try to avoid such situations. |
| Long-Term Orientation (vs Short-Term Orientation*) | Values oriented towards future rewards, perseverance, and thrift, which are related to 'saving' as opposed to 'spending'. |
| Power Distance (high vs low) | A degree to which inequality and unequal distributions of power between parties are accepted. |
| Success Drivenness (vs Quality-of-Life Orientation) | A societal value for achievement, and material rewards for success. |

\*Associated with higher fidelity scores in the Recovery College operational model.

This study builds on prior work that compared how RCs are introduced to the public in England and Japan by analysing their promotional texts (21). That earlier study found that the way RCs are framed aligned with key cultural characteristics (namely Individualism vs. Collectivism and Short-Term vs. Long-Term Orientation) highlighting how RCs are presented to appeal to the public values. For instance, the promotional texts in England emphasised self-management and skill acquisition, reflecting an individualistic and short-term orientation, while the texts in Japan emphasised group life-long learning, consistent with collectivism and long-term orientation.

In the current study, the Netherlands is introduced as a third comparison country. Dutch culture is broadly similar to English culture in five dimensions, but it uniquely stands out in its strong emphasis on quality of life (Quality-of-Life Orientation rather than Success Drivenness) (26): a value that stands in contrast with the success-driven English culture and is virtually absent in Japan, where a high drive for achievement dominates (27). Japanese culture contrasts even more strongly with Dutch culture across multiple dimensions, such as Individualism vs. Collectivism, and Short-Term Orientation vs. Long-Term Orientation. Table 2 presents these cultural metrics for the three countries (27). These distinctions are important given that all three countries actively implement the RC model, with 88 RCs in England, 25 in the Netherlands, and 15 in Japan. Therefore, comparing these three countries can yield meaningful cross-cultural findings.

**Table 2.** Metrics for the six cultural characteristics in the Netherlands, England, and Japan.

| Cultural<br>Characteristic (rows)<br>Country (columns) | Individualism | Indulgence | Uncertainty<br>Avoidance | Long-Term<br>Orientation | Power<br>Distance | Success<br>Drivenness |
| --- | --- | --- | --- | --- | --- | --- |
| Netherlands | 80 | 68 | 53 | 67 | 38 | 14 |
| UK | 89 | 69 | 35 | 51 | 35 | 66 |
| Japan | 46 | 42 | 92 | 88 | 54 | 95 |

The recovery movement has made substantial progress in the Netherlands over the past two decades (28, 29). Driven by both international recovery movements and local influential initiatives such as the Hearing Voices movement (30), the Dutch mental healthcare system has gradually shifted from a purely medical model toward a more holistic approach. This transformation is reflected in national care standards that prioritise connectedness, hope, identity, meaning, and empowerment: core elements of the well-established CHIME recovery framework (11, 31). Lived experience has become a recognised form of expertise, with trained experts now being part of multidisciplinary teams. In recent years, RCs have also gained increasing traction and public support across the country (32) (33). These developments signal the growing commitment to personal and social recovery on an institutional level, moving beyond a sole focus on symptom reduction.

This study aimed to (a) identify the textual emphases in how RCs are presented to the public in the Netherlands, and (b) explore how the Quality-of-Life Orientation is reflected in the promotional texts in the Netherlands through comparison with those in England and Japan.

## Methods

### Design

We conducted a qualitative and quantitative text-based analysis of publicly available online promotional texts from Recovery Colleges (RCs) in the Netherlands to examine how recovery and RCs are characterised, addressing Aim (a). To address Aim (b), these findings were compared with previously reported data from RCs in Japan and England, interpreted using Hofstede’s Cultural Dimensions Theory (21). Ethical approval was not required, as all texts were publicly available.

### Data

Promotional texts were collected from the websites of 25 RCs in the Netherlands by MA and RR (23 May 2025). The texts on the RC webpages explaining what RCs and recovery are, were first identified by MA, which were then reviewed by RR. All URLs are available on the Open Science Framework at https://osf.io/9f7pv/. Dutch texts were translated into English by MA, a Dutch first-language speaker versed in RC terminology and practice and fluent in English.

### Data Analysis

We performed a corpus-based discourse analysis (34), which involves the software-assisted examination of large collections of digitised texts of naturally occurring language (“corpus”), using the online software Sketch Engine (35). This analysis was structured around seven themes identified in the previous England-Japan study (21): Personal learning, Place for wellbeing, Recovery, Education, Community, Service users, and Mental illness experience. Textual analysis was performed using three common corpus tools to allow for quantitative comparisons: Wordlist, Word Sketch and Keywords. Interpretation of textual patterns was guided by Hofstede’s Cultural Dimensions Theory (25) and the cross-cultural framework for critical discourse analysis in mental health recovery research (36), a novel analytical model designed to contextualise discourse within sociocultural and recovery-oriented paradigms.

### Wordlist

Wordlist retrieves lists of words from the datasets with their raw frequency (RaF: number of occurrences). Relative frequency (ReF) was calculated to allow comparisons between the datasets. ReF is the ratio between the RaF of a word and the total number of words in the dataset, expressed per 1,000 words (1,000 was chosen as being close to the dataset sizes: 2,976 words in the Netherlands, 22,014 in England, and 813 in Japan). We examined the top-ten verbs (processes), nouns (entities), and adjectives (descriptive terms). Words that (a) show high variation in use in the datasets, and (b) could not be grouped within one of the main themes were excluded (see Table 4, section results). For instance, the verbs ‘have’ and ‘be’ were excluded since they may refer to service users, the RCs, any activity, or may be used grammatically (e.g. ‘have’ in the present perfect tense).

### Word sketch

Word Sketch extracts “collocates”, words co-occurring with a focus word (“recovery”) with a frequency higher than chance. Examining collocates for “recovery” shows patterns of use of the word and meanings associated with it.

### Keywords

Keywords are words statistically significant in a dataset as compared to another dataset of reference (37). Keywords reflect the uniqueness of a dataset and make it possible to identify key themes. To allow for comparison between the keywords of each dataset (Dutch RCs, English RCs, Japanese RCs), the general corpus English Web enTenTen21 (38) (available in Sketch Engine) was used as reference corpus for comparison. The enTenTen21 includes over 52 billion words of online texts covering a range of topics (e.g. health, law) extracted from websites of domains of countries with English as official language. We examined the top 50 keywords and focused on lexical words (e.g., nouns, verbs), which are content-informative compared with grammatical words such as prepositions. In Sketch Engine, keyness score (i.e. the measure that indicates how a word is statistically distinctive within one dataset compared to the reference dataset) is calculated as follows:

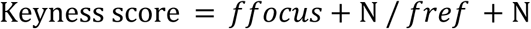

*f_focus_*is the normalised (per million) frequency of the word in the focus dataset (corpus), *f_ref_*is the normalised frequency of the same word in the reference corpus, and N the smoothing parameter (for this keyword extraction, N=100)(35).

The concordance tool was used to examine the context of use of high frequency words and keywords to identify patterns of use and themes. All methodological terms are appended in S1 Supporting Information.

## Results

### Retrieved Data

Dutch text data comprised 3,445 words after translation into English. Existing text datasets from RCs in England and in Japan comprised a total of 22,014 and 813 words respectively, the latter after translation into English. The 25 Dutch RCs are shown in Table 3.

**Table 3.** Dutch RCs (n=25) with wordcounts for each college.

| **RCs in the Netherlands** | **Words** |
| --- | --- |
| Leger des Heils Herstelacademie | 119 |
| Herstelacademie Haarlem en Meer | 114 |
| Herstelacademie GGz Noord-Holland-Noord | 154 |
| De Herstelacademies in Rotterdam | 197 |
| Enik Recovery College | 137 |
| FAMEUS | 141 |
| Herstelacademie Doel Delfland | 167 |
| Herstelacademies Reinier van Arkel (Bommelerwaard, de Stenen Hut, de Stijl) | 163 |
| PEERpoint | 138 |
| Sedna | 356 |
| Mens door mens | 69 |
| Herstelacademie Venlo/Roermond | 83 |
| Korak herstelacademie | 113 |
| Stjoer | 142 |
| Herstelacademie Thuis | 125 |
| Herstelruimte Emmen | 53 |
| Nexus | 92 |
| Herstelacademie Stadskamer | 181 |
| Steunpunt Zelfregie & Herstel | 105 |
| Herstelacademie voor jongvolwassenen (Reakt) | 104 |
| Herstelbureau van HVO-Querido | 136 |
| Recovery College Eindhoven | 87 |
| Herstelacademie de Kiem | 125 |
| Ixta Noa | 202 |
| HerstelXL | 142 |
| **Total** | **3,445** |

### Overview

Wordlist (frequency analysis) revealed that the “Recovery” theme was most prominent in the Dutch dataset (the Word Sketch is presented in the theme-level findings). In contrast, explicit references to “Education” were notably absent from the top-ten most frequent nouns (entities), verbs (processes), or adjectives (descriptive words) (Table 4). This stands in contrast to the English RC dataset, where “Education” emerged as the most dominant theme.

**Table 4.**
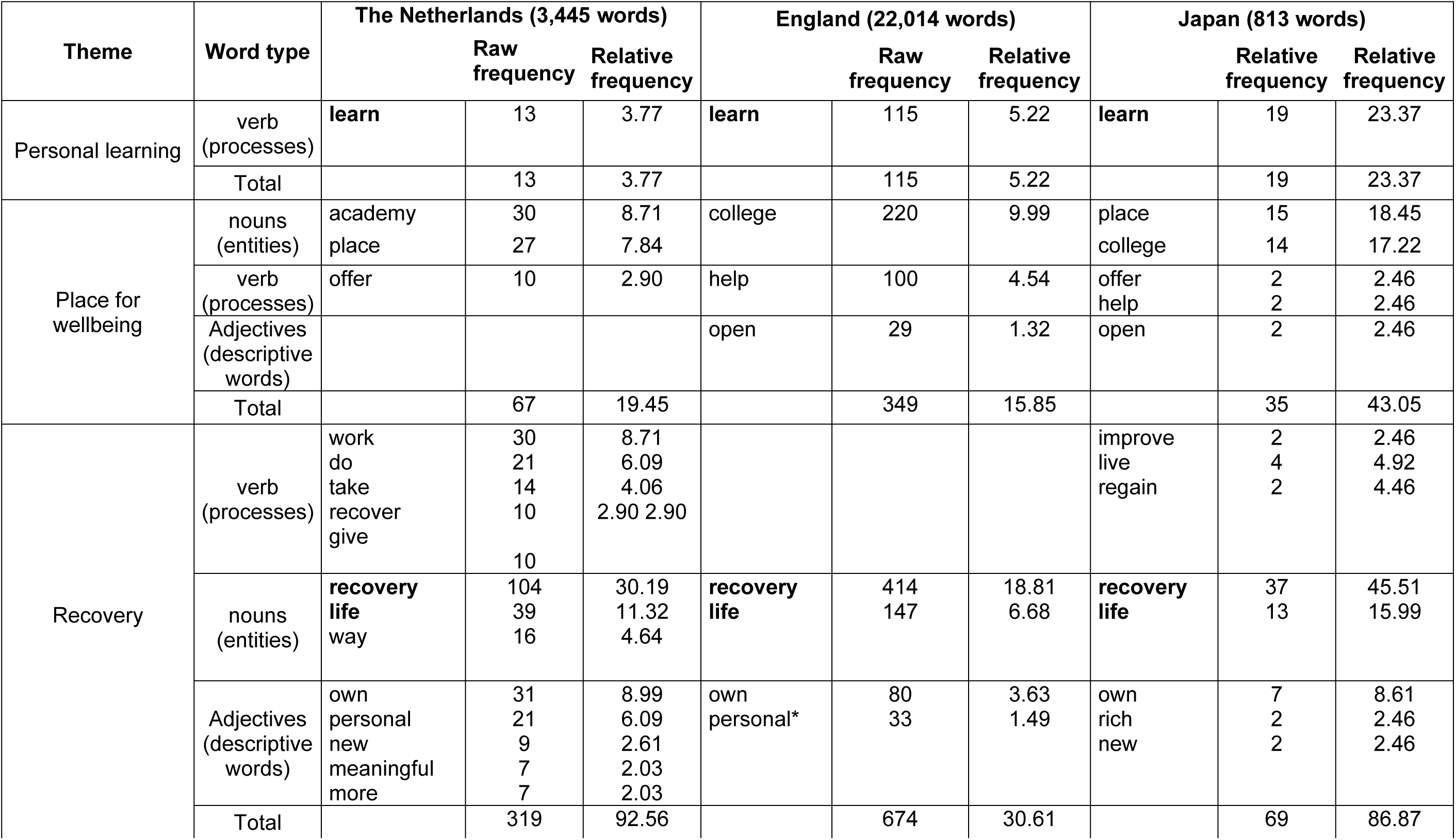
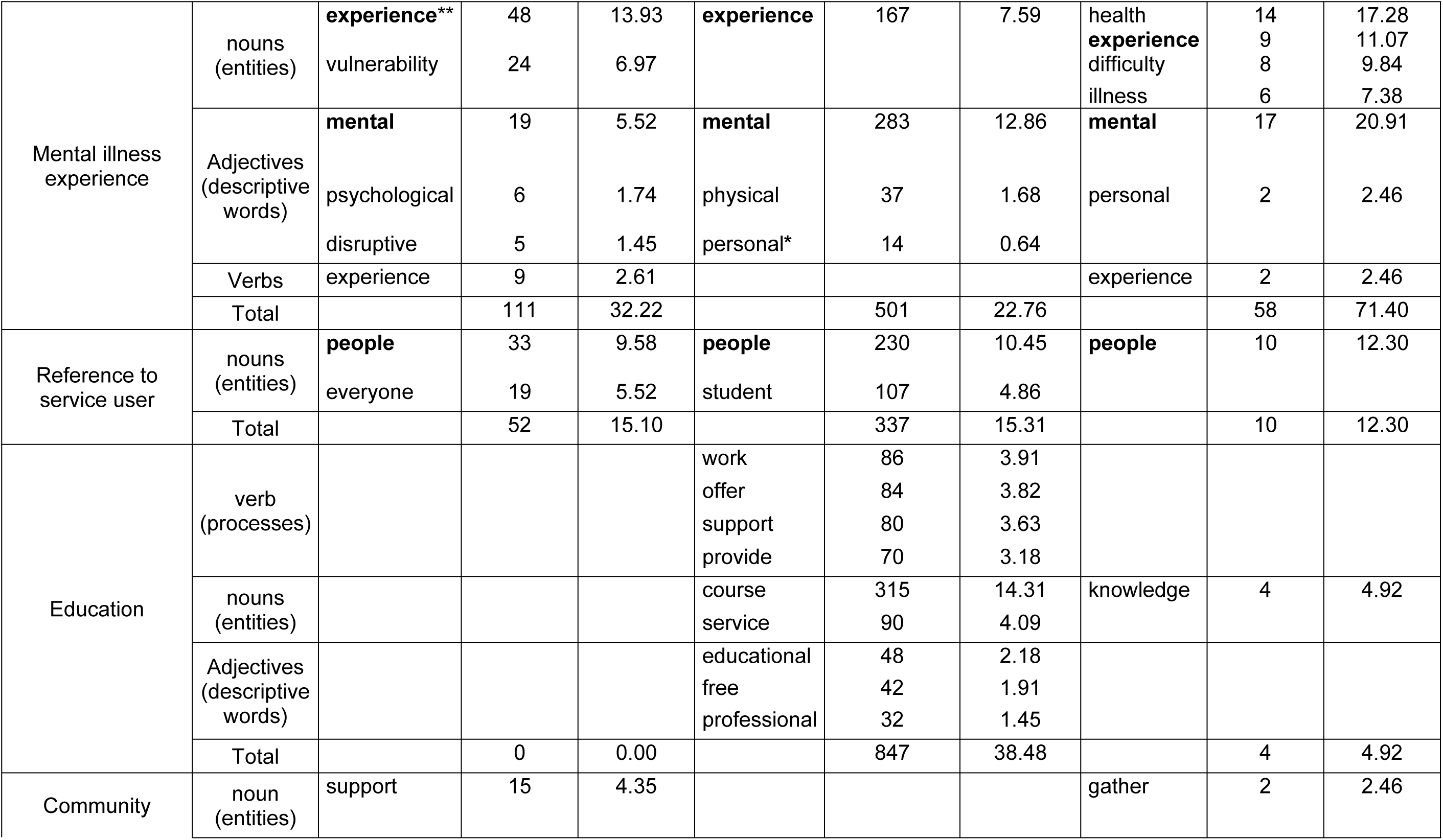
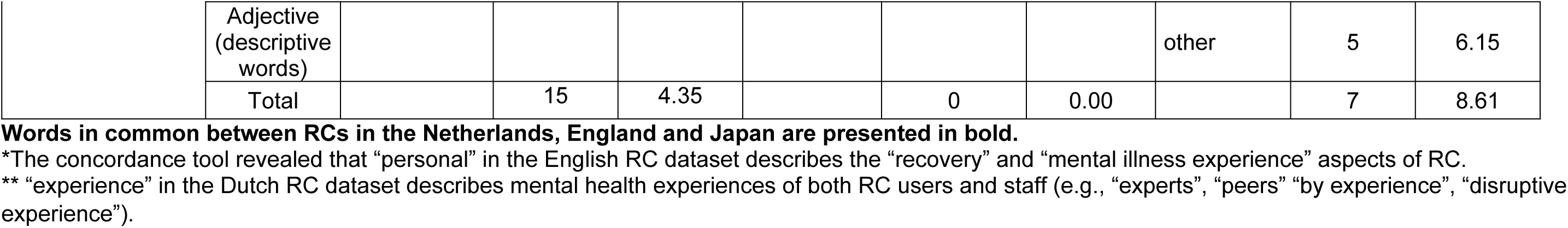
Wordlists with raw and relative frequencies for top-ten verbs, nouns and adjectives in the Dutch, English and Japanese RCs.

Keyword analysis revealed that “Education” and “Personal learning” were defining themes in the English RC texts yet were almost entirely absent from the Dutch materials (Table 5). Conversely, “Recovery” and “Mental illness experience” were more prominent in the Dutch and Japanese RC datasets, suggesting a shared thematic focus distinct from the English texts.

**Table 5.** Keywords for the Dutch, English and Japanese RC datasets compared to English enTenTen21.

|  | The Netherlands (2,976 words) | England (22,014 words) | Japan (813 words) |
| --- | --- | --- | --- |
| Theme | Single (raw frequency / keyness score) | Single (raw frequency / keyness score)<br>Total keyness score | Single (raw frequency / keyness score) |
| Personal learning | learn (13/7.75) | skill-management (29/13.25), skill (73/12.78), learn (113/11.29), learning (28/7.80), knowledge (39/7.18) | learn (19/47.38), knowledge (4/18.02), expertise (2/16.81), proactive (1/11.10), wellbeing (1/10.82) |
| Place for wellbeing | academy (30/52.768), Mens (4/10.84), Ixta (4/11.07), Noa (4/11.03) place (27/9.66), Herstelxl (3/8.55), Peerpoint (3/8.55), Enik (3/8.55), Ggz (3/8.55), meeting (8/7.86) | college (212/31.23), academy (26/8.32), NHS, (18/7.75) | college (14/52.01), welfare (3/26.87), place (15/22.81), provider (2/12.87), professional (3/11.83), RC (1/11.17), accumulate (1/10.42) |
| Recovery | recovery (104/173.39), recover (10/18.23), personal (21/19.88), self-direction (5/13.58), meaningful (7/17.46), life (39/13.91), cope (5/11.73), healing (5/11.25), strength (6/9.38), meaning (5/9.06), fulfill (4/8.87), hopeful (3/7.96), feeling (5/7.89), process (15/8.20), regain (3/7.83), balance (5/7.67) | recovery (402/113.69), wellbeing (137/54.49), journey (51/14.09), personal (60/9.81), life (145/8.81), hope (65/8.46), meaningful (21/8.26), empower (18/7.29) | recovery (37/264.19), wisdom (4/35.52), regain (2/20.68), life (13/19.82), meaning (2/15.07), rich (2/13.01), concept (2/11.42), reconstruct (1/11.27), satisfying (1/11.18), functioning (1/11.13), livelihood (1/11.06), hopeful (1/10.98), enrich (1/10.72), unique (2/10.65), refer (2/10.38) |
| Mental illness experience | vulnerability (24/53.63), mental (19/29.63), disruption (9/21.32), disruptive (5/13.01), illness (7/14.21), psychological (6/13.31), limitation (5/10.66), life-disrupting (3/8.55), | mental (265/69.13), health (294/26.48), illness (33/11.49), challenge (60/9.27), physical (36/7.78) | mental (17/111.96), difficulty (8/60.66), illness (6/50.19), health (14/31.89), disability (3/24.44), symptom (2/15.49), living (2/15.21), challenge (3/11.65), prognosis (1/11.50), lessen (1/11.23), psychiatric (1/11.09), hardship (1/11.01) |
| Reference to service user | everyone (19/17.95), participant (6/9.34) | people (220/7.85), student (106/7.55) |  |
| Education | experience (57/27.19), peer (9/18.2), expert (15/19.46), organise (4/8.94), participate (7/10.07), volunteer (6/9.57), expertise (4/8.23), training (8/7.49) | course (298/27.90), peer (54/18.47), co-production (34/15.33), tutor (38/15.12), trainer (38/14.73), education (47/13.53), workshop (43/12.04), professional (76/11.83), partnership (39/10.80), opportunity (74/10.32), co-produce (20/9.41), deliver (47/9.21), expert (40/9.06), session (38/8.46), aim (36/8.14), staff (47/7.91), attend (35/7.76), organisation (26/7.68), Trust (34/7.43), offer (89/7.39), range (51/7.19), recognise (19/7.15) |  |
| Community | recognition (5/9.87), together (13/9.54), support (23/8.89) | carer (43/18.39), support (137/8.96) |  |

Keywords not coded by theme and references to locations of specific RCs have not been included (e.g., "Oxfordshire"). "Mens", "Peerpoint" and "Enik" refer to specific recovery collages in the Netherlands.

### Theme-level Findings

#### Personal learning

This theme emphasises knowledge development, skills acquisition and development of self-management. The keyword “learn” (treated as a lemma, i.e., its standard dictionary form that includes variations such as “learned” and “learning”) was the only term recurrently associated with this theme across all three datasets (Tables 4-5). Concordances for “learn” were examined to identify further differences (Table 6), which includes three random concordances for each one of the datasets.

**Table 6.** Concordances for “learn”.

| Country | Examples: “learn” (lemma*) |  |
| --- | --- | --- |
| The Netherlands | 1 | Working on recovery is <u>learning</u> to cope with your mental vulnerability. |
|  | 2 | ... gaining more insight into your personal experiences and disruption, <u>learning</u> to cope better with your vulnerabilities and taking control of your life again. |
|  | 3 | ... a recovery academy: a <u>learning</u> environment and meeting place for people who have personal experience of disruption... |
| England | 4 | ... runs a wide range of courses, workshops and activities to help you better understand your mental health and <u>learn</u> strategies to manage it better. |
|  | 5 | The ethos is to increase students’ knowledge and skills and help them take control by <u>learning</u> self-management strategies to help their mental health and wellbeing. |
|  | 6 | It uses feedback from students to monitor the success of the individual’s <u>learning</u> experience and their personal goals for gaining hope, opportunity and control of their lives. |
| Japan | 7 | At Recovery College, we believe in <u>learning</u> “together” by bringing our perspectives and experiences to the table. |
|  | 8 | Recovery is about people from all walks of life <u>learning</u> together about regaining their unique way of going about their lives. |
|  | 9 | ...people with difficulties in living, family members, supporters, and community members gather to <u>learn</u> together about living life on their own terms. |

RCs in Japan emphasise learning as a communal practice, conflating community experience with learning (Examples 7-9 (E7-9), also emphasis on ‘learn[ing] from each other’, 5 times). Although RCs in England also refer to communal learning (21), the emphasis is on individual learning, acquisition of skills and control (“self-management”) and the range of the educational offer (E4-6). The Netherlands RCs also emphasise individual learning in comparison to community (E1-3). However, instead of focusing on the educational nature of the RCs, the emphasis is on identifying (individual) learning with coping with one’s experiences and acquiring control (E1-2).

#### Place for wellbeing

The theme “Place for wellbeing” was particularly prominent in the Japanese RC dataset (Table 7).

**Table 7.**
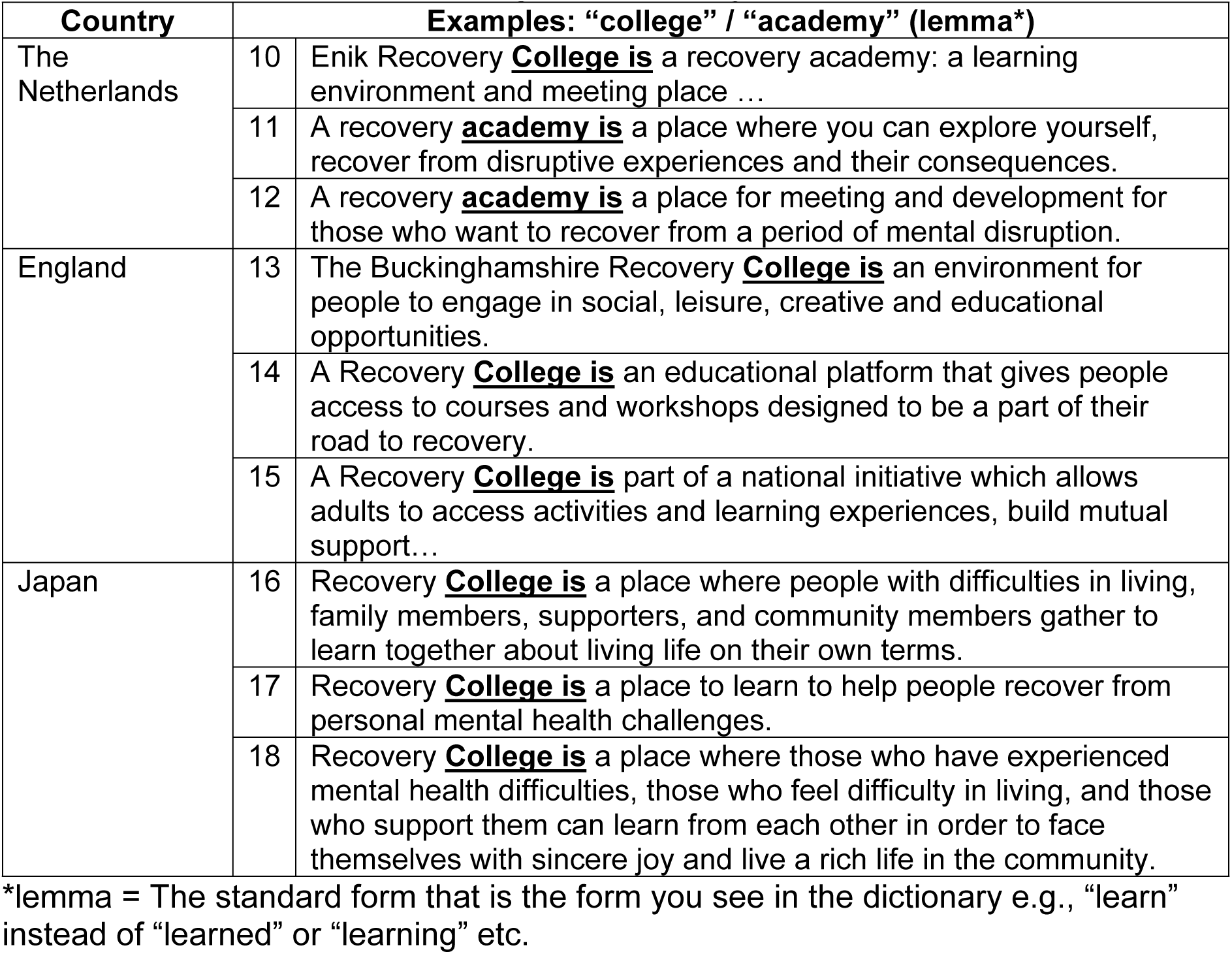
Concordances for “college” / “academy”.

| Country | Examples: “college” / “academy” (lemma*) |  |
| --- | --- | --- |
| The Netherlands | 10 | Enik Recovery **College is** a recovery academy: a learning environment and meeting place ... |
|  | 11 | A recovery **academy is** a place where you can explore yourself, recover from disruptive experiences and their consequences. |
|  | 12 | A recovery **academy is** a place for meeting and development for those who want to recover from a period of mental disruption. |
| England | 13 | The Buckinghamshire Recovery **College is** an environment for people to engage in social, leisure, creative and educational opportunities. |
|  | 14 | A Recovery **College is** an educational platform that gives people access to courses and workshops designed to be a part of their road to recovery. |
|  | 15 | A Recovery **College is** part of a national initiative which allows adults to access activities and learning experiences, build mutual support... |
| Japan | 16 | Recovery **College is** a place where people with difficulties in living, family members, supporters, and community members gather to learn together about living life on their own terms. |
|  | 17 | Recovery **College is** a place to learn to help people recover from personal mental health challenges. |
|  | 18 | Recovery **College is** a place where those who have experienced mental health difficulties, those who feel difficulty in living, and those who support them can learn from each other in order to face themselves with sincere joy and live a rich life in the community. |

\*lemma = The standard form that is the form you see in the dictionary e.g., “learn” instead of “learned” or “learning” etc.

References to RCs varied across the three national contexts: while the Japanese and English datasets frequently referenced the term “college” (Table 2), the Dutch dataset predominantly used “academy”, with only three instances referring to a specific RC that includes “college” in its formal name (Table 5, E10). Table 7 presents three randomly selected concordance lines per dataset for the phrase “college/academy is”, illustrating how the concept is framed in different linguistic and cultural contexts.

Across all three datasets, college/academy is consistently constructed as a site for recovery-oriented learning (E10, E14–15, E16–18) and communal experience. However, subtle thematic emphases emerge. In the English dataset, the college is explicitly positioned as an educational setting (E14–15). In contrast, the Dutch dataset highlights academy as a space for personal development within a community (E11– 12). The Japanese dataset, meanwhile, frames college primarily as a communal space that supports wellbeing (E16, E18), reflecting its emphasis on social connectedness as a key component of recovery.

#### Recovery

The theme “Recovery” was particularly prominent in both the Dutch and Japanese RC datasets (Table 8). However, the contextual framing of recovery differs across the three countries, as illustrated in the Word Sketch visualisations (Fig 1).

**Fig 1.**
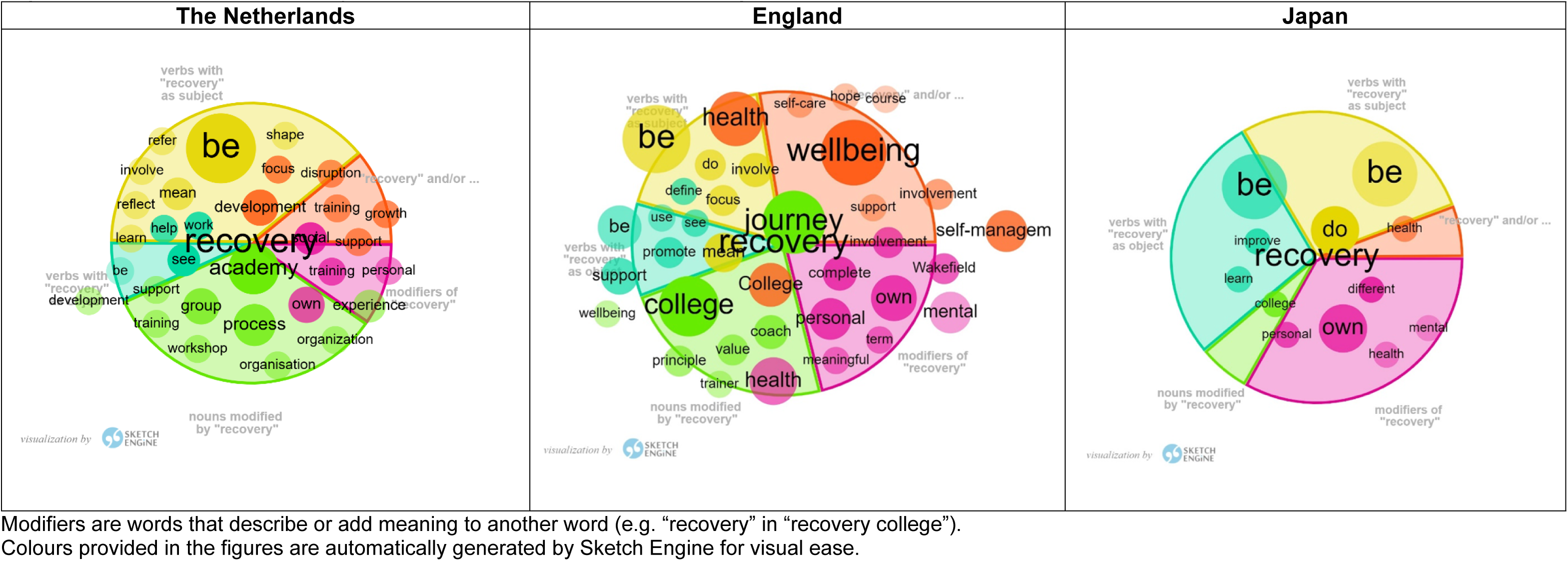
**Collocates of “recovery” for RCs in the Netherlands, England and Japan RCs**

**Table 8.** Concordances for “recovery”.

| Country | Examples: “recovery” (lemma*) |  |
| --- | --- | --- |
| The Netherlands | 19 | By **recovery**, we mean gradually managing to arrange your life in a way that you like. |
|  | 20 | **<u>Recovery</u>** is not about 'curing' your 'illness', but about gaining more insight into your personal experiences and disruption, learning to cope better with your vulnerabilities and taking control of your life again. |
|  | 21 | **<u>Recovery</u>** is a unique and deeply personal process in which a person changes their attitudes, values, feelings, goals, skills, and roles. |
| England | 22 | **<u>Recovery</u>** is a personal journey. |
|  | 23 | **<u>Recovery</u>** does not always refer to the process of complete recovery from a mental or physical health condition. It is about recovering a life worth living ... |
|  | 24 | ... **<u>recovery</u>** is the process of rebuilding a satisfying, hopeful and contributing life with a diagnosis of mental health problems. |
| Japan | 25 | The conclusion of **<u>Recovery</u>** was that if there are 100 people, there are 100 different Recoveries... |
|  | 26 | ... **<u>recovery</u>** does not happen alone, but in a relationship with people you trust. |
|  | 27 | **<u>Recovery</u>** is about people from all walks of life learning together about regaining their unique way of going about their lives. |

In the English dataset, recovery is predominantly conceptualised as an individualised process associated with terms such as “personal”, “own”, “self-management”, “wellbeing”, and “journey”, and is often framed as something to be achieved through education (e.g., “coach”, “trainer”).

While “personal” also emerges as a key descriptor of recovery in the Dutch and Japanese datasets, other dimensions are foregrounded. The Dutch dataset includes references to educational formats such as “workshop”, “training”, and “development”, but places greater emphasis on community-oriented concepts including “group”, “social”, “support”, and “help”. In the Japanese dataset, the smaller volume of text yielded fewer collocates overall; however, among those retrieved, recovery was notably associated with “mental health” improvement and the notion of being “different”, suggesting a nuanced view tied to identity and societal positioning.

To gain deeper insight, concordance lines for the term “recovery” were examined, excluding references to “recovery college/academy” and institutional names. Table 6 presents three randomly selected concordances per dataset.

Across all three countries, RCs depict recovery as a process of regaining meaning and hope in life (E20, E23–24, E27), and as a deeply personal journey (E21–22, E25). However, specific emphases diverge. Japanese RCs highlight the role of community in supporting recovery (E26 and (21)), whereas English and Dutch RCs place stronger emphasis on individual growth, self-direction, and taking control of one’s life (Table 8, E20).

#### Education

The theme “Education” was most prominent in the English RC dataset, and to a much lesser extent in the Dutch dataset (Table 5). In contrast, the Japanese RC texts did not feature any of the education-related keywords observed in the English and Dutch datasets (Table 5), nor did they include any of the high-frequency educational terms identified in the English corpus (Table 4).

Table 9 presents three randomly selected concordances for the term “course” from the English and Dutch datasets, along with the only two concordances retrieved from the Japanese dataset.

**Table 9.**
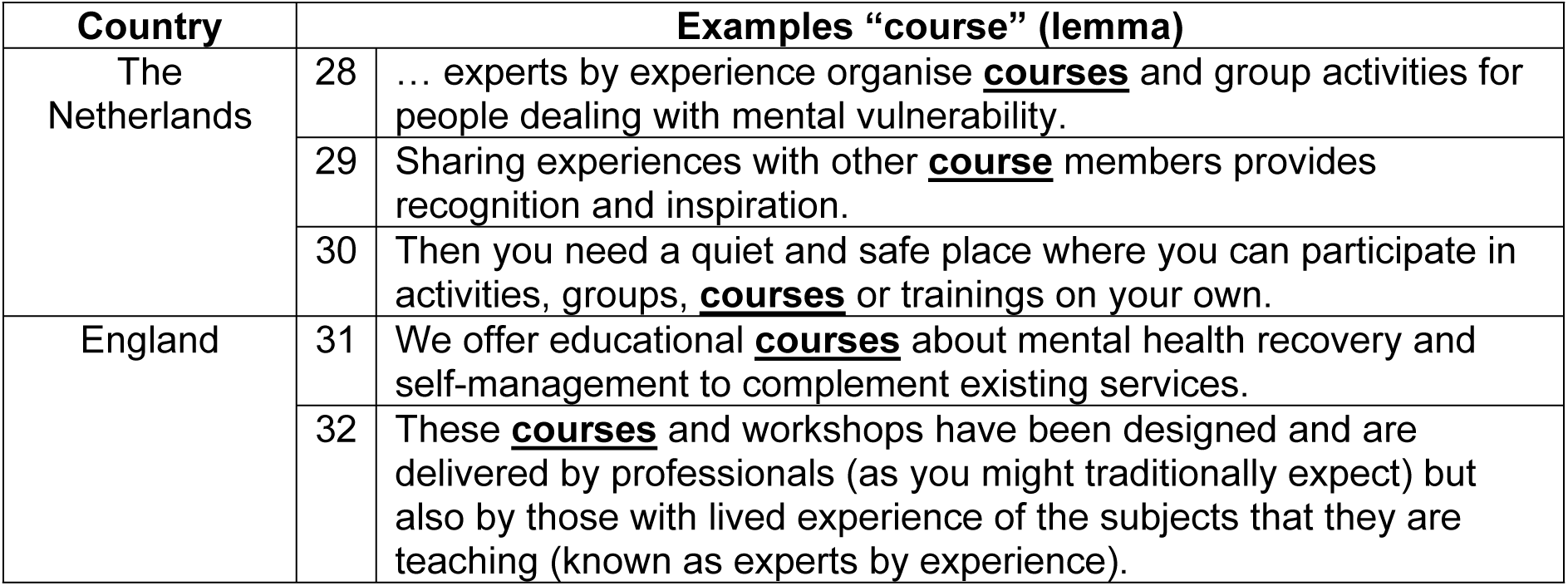
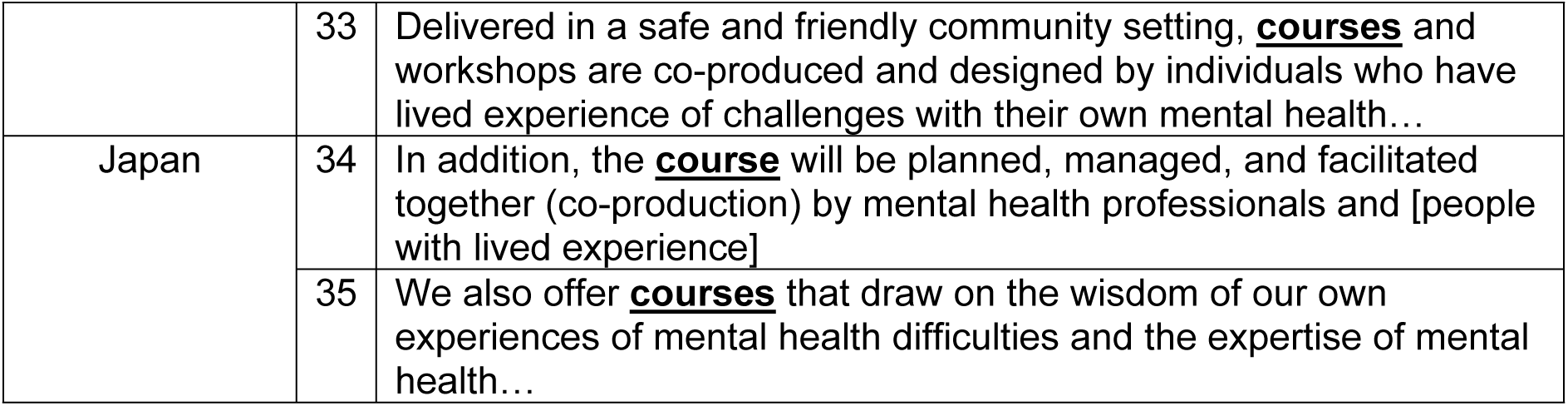
Concordances for “course”.

In both the Japanese (E34-35) and English (E32-33) RCs, the co-designed nature of courses is emphasised. English RCs additionally highlight the specific content and skills to be acquired through these courses (E31). In the Dutch dataset, courses are mentioned primarily as examples of available activities, with a greater focus on their communal function rather than on individual learning or formal education (E28-30).

#### Community

The theme “Community” is primarily used in the English RC dataset to refer to external partnerships (e.g., “community organisations” and “community partners”), or to specific communities receiving support (Table 10, E39–40), although it is also referenced as a component of the recovery process (E41). In contrast, Japanese RCs frame community as integral to personal learning in recovery (Table 6) and position the RC itself as a communal space (Table 7). Similarly, Dutch RCs are portrayed as spaces of togetherness, frequently described as a “meeting place” (five instances; Table 7).

**Table 10.** Concordances for “community”.

| Country | Examples: “course” (lemma) |  |
| --- | --- | --- |
| The Netherlands | 36 | A fulfilling and hopeful life, in which you can make a meaningful contribution to **society**, is also possible for you... |
|  | 37 | This new view of health is about self-direction, taking responsibility, adapting yourself and participating in **society**. |
|  | 38 | ... projects and activities where people with mental and/or social vulnerability can shape their participation in **society** without stigma. |
| England | 39 | ...provides recovery-focused education courses that are accessible to everyone in our local **communities**. |
|  | 40 | We work alongside and support students, volunteers, **community** organisations and healthcare professionals... |
|  | 41 | Recovery is holistic and embraces the whole person, including their mind, body, spirit and **community**. |
| Japan | 42 | RC is a learning **community** where anyone can participate... |
|  | 43 | ...where people with illnesses and their supporters can learn from each other the knowledge they need to live well in the **community**... |
|  | 44 | ... can learn from each other in order to face themselves with sincere joy and live a rich life in the **community**. |

While the English and Japanese RC datasets include explicit references to “community”, this term is absent in the Dutch RC texts. Instead, references to “society” appear (three instances; Table 7), potentially reflecting translation or cultural nuances. In the Japanese RC dataset, concordances for “community” (E42–44) reflect its role in fostering connectedness and recovery. Meanwhile, references to “society” (E36–38) in the Dutch RCs emphasise the goal of recovery as the ability to live well in society and to make meaningful contributions.

#### Service users

The theme “service users” is the least emphasised across all three datasets. In each country, RCs mostly refer to service users using the general noun “people”, often accompanied by mentions of their mental health experiences, especially in RCs in Japan (E51-52) and the Netherlands (E45-46). RCs in Japan exclusively use “people” to refer to service users (Table 11), and include references to their different backgrounds (E53) emphasising RCs’ inclusivity. RCs in the Netherlands also emphasise togetherness: both through references to “people” (E47) and through the inclusive pronoun “everyone” (Table 11, e.g. “Everyone is welcome…”, “Everyone in the group has certain similarities…”). Instead, RCs in England emphasise the educational perspective. Service users are referred to as “people” and “student” (Table 11). While “people” is used more than twice as often as “student” (Table 11), these mentions often hint at the educational role of RCs and suggest that service users are also framed as learners within these settings.

**Table 11.** Concordances for “people”.

| Country | Examples: “people” (lemma) |  |
| --- | --- | --- |
| The Netherlands | 45 | ... are places for and by <u>people</u> with psychological vulnerability. |
|  | 46 | Nexus is a place where <u>people</u> with mental vulnerability can develop... |
|  | 47 | You experience this process [recovery] together with <u>people</u> you can fall back on. |
| England | 48 | ... by recognising resourcefulness, talents and skills, <u>people</u> become experts in their own health and wellbeing... |
|  | 49 | We create a safe place where <u>people</u> can come together to learn ways to live healthier... |
|  | 50 | Provide a peer led, peer delivered education and support service where <u>people</u> can learn from each other's insights... |
| Japan | 51 | Recovery College is a place where <u>people</u> with difficulties in living, family members... |
|  | 52 | Recovery College is expanding as a place where <u>people</u> with illnesses and their supporters... |
|  | 53 | Recovery is about <u>people</u> from all walks of life learning together ... |

#### Mental illness experience

The theme “mental illness experience” is relevant in all three datasets, reflecting the topic of the promotional texts (Tables 4 and 5). However, mental illness is accounted for differently. Dutch RCs emphasise “vulnerability” and life disruption (“disruption”, “disruptive”, “life-disrupting”), and include explicit references to addiction (Table 5 and E54-56 in Table 12). References to addiction are also observed in RCs in England, which feature 14 references to “substance misuse problems” and three references to “addiction”, but not in RCs in Japan. RCs in England and Japan emphasise the negative impact of mental health problems (Table 5; e.g. English RCs refer to “challenges”, “difficulties” and “problems”, with 30, 13 and 8 occurrences respectively). RCs in England and Japan RCs adopt a comparatively more medicalised discourse than RCs in the Netherlands, and include references to “mental illness”/“mental or physical illness” (21 occurrences in England and one in Japan), to “mental ill health” and “mental health diagnosis” (seven and one occurrences respectively in England), and to “mental disorder” (Japan, one occurrence) (Table 12, E57-62). RCs in Japan also refer to “disability” and those in England to “physical” health/illness (Table 5).

**Table 12.** Concordances for “mental”.

| Country | Examples: “mental” (lemma) |  |
| --- | --- | --- |
| The Netherlands | 54 | Working on recovery is learning to cope with your <u>mental</u> vulnerability. |
|  | 55 | That could be recovery from addiction, <u>mental</u> vulnerability and/or other unpleasant experiences. |
|  | 56 | ... recovery is not about healing, but about discovering your potential for a fulfilling life with <u>mental</u> vulnerability. |
| England | 57 | ... anyone wishing to improve their mental wellbeing or recovering from a period of <u>mental</u> ill-health. |
|  | 58 | Being diagnosed with <u>mental</u> illness can be very frightening. |
|  | 59 | You don't need to be referred to the college, have a <u>mental</u> health diagnosis... |
| Japan | 60 | ... those who have experience of <u>mental</u> illness who are in the process of recovery ... |
|  | 61 | Recovery here does not simply refer to improvement in <u>mental</u> symptoms or functioning... |
|  | 62 | ... based on discussions and social movements among people with <u>mental</u> disorders. |

### Summary of Findings

To address Aim (a) regarding the textual emphases in how RCs are presented to the public in the Netherlands, our analysis identified “recovery” as the most prominent theme in Dutch RC promotional texts. These texts frequently referred to personal growth, experiences of mental illness, and communal support. Unlike the English dataset, educational terms were largely absent; instead, Dutch RCs were portrayed as “academies” or “meeting places,” suggesting spaces for reflection, interaction, and recovery. The emphasis on terms such as “society” rather than “community” indicates a framing of RCs as socially integrative environments rather than closed support networks.

To address Aim (b) regarding how the Quality-of-Life Orientation is reflected in the Dutch RC texts through comparison with those in England and Japan, we observed distinct patterns. English RCs tended to frame recovery as an individualised, educational journey, strongly linked to self-management and structured learning. Japanese RCs highlighted collective and relational aspects, portraying recovery as grounded in inclusive learning and mutual support. Dutch RCs occupied a middle ground: they emphasised personal development and empowerment, but within a socially embedded context. In addition, Dutch RCs frequently referenced the idea of recovery in everyday life, highlighting the importance of practical coping, daily functioning, and social participation. These emphases on togetherness, societal contribution, and daily life functioning resonate with the characteristics of a Quality-of-Life Orientation, which prioritises wellbeing, social connection, and meaningful engagement.

## Discussion

This study investigated how RCs in the Netherlands are presented to the public, and compared the data with English and Japanese RCs. A total of 3,445 words from 25 RCs in the Netherlands were analysed using a corpus linguistic approach. Words such as “recovery” and “society” were emphasised in the promotional texts of Dutch RCs. Comparing with the promotional texts from England and Japan, the Dutch texts foregrounded togetherness, societal contribution, and daily life functioning, indicating the impact of the Quality-of-Life Orientation.

Regarding Aim (a), the Dutch RC texts most prominently featured the theme of “recovery”, conceptualised as a personal yet socially contextualised process. While references to mental illness experiences were frequent, explicit mentions of education were largely absent, in contrast to English RCs. Instead, Dutch RCs presented themselves as “academies” or “meeting places”, indicating spaces for communal interaction, reflection, and support. The absence of the term “community” and preference for “society” further indicated an orientation toward broader social integration, rather than enclosed communal identity. This aligns with the Dutch mental health policy landscape, which increasingly centres societal participation and relational recovery (11, 39) (40), and with recent literature describing RCs as vehicles for restoring social roles and connectedness rather than providing education for self-management skill acquisition (32, 33).

Regarding Aim (b), the Netherlands-England-Japan comparison revealed culturally distinctive constructions of recovery, with Dutch RCs reflecting a unique textual orientation grounded in everyday life and social connection (41). English RCs framed recovery as an individualised, skills-based journey embedded in a structured educational framework, consistent with England’s Success-Driven and Short-Term Orientation values (26). Japanese RCs, by contrast, described recovery as a communal, long-term process of shared meaning-making, aligning with Collectivist and Long-Term Orientation values (21).

Dutch RCs represented a midpoint between these cultural poles: while recovery was presented as a personal journey, it was firmly situated within social relationships and the realities of daily life. Learning was framed not in terms of acquiring credentials or professional success, but as regaining agency and coping with lived experiences. This pattern aligns with how recovery is commonly framed in the Netherlands, often as *herstel in het dagelijks leven* (“recovery in daily life”), foregrounding personal agency, relational embeddedness, and engagement in ordinary societal roles (42–44). These emphases also reflect broader Dutch cultural values, such as wellbeing, moderation, and equality, that shape how recovery is communicated and enacted in practice. Words such as “meeting place”, “self-direction”, and “participation in society” embody this ethos. In mental health settings, these values are operationalised through principles such as normalisation and citizenship, where recovery is understood as meaningful participation in everyday life (43, 45, 46).

A major strength of this study lies in its cross-cultural and linguistic comparative design, drawing on a substantial corpus of RC promotional texts across three culturally diverse settings. The use of corpus-based discourse analysis, supported by robust computational tools (Sketch Engine), enabled a systematic, fine-grained comparison of lexical patterns, thematic prominence, and socio-cultural framing. Moreover, the integration of Hofstede’s Cultural Dimensions Theory and a novel cross-cultural framework for critical discourse analysis lends conceptual depth and theoretical rigour to our interpretations. By focusing on public-facing texts, this study also captures the cultural messaging strategies that may shape public understanding and engagement with RCs.

Several limitations should be noted. First, the size of the Japanese dataset was considerably smaller than those of the Netherlands and England, which may have constrained the breadth of linguistic features identified. Second, translation effects from Dutch and Japanese to English, may have influenced the lexical and semantic nuances detected, despite the translators’ expertise. Third, as this study focused exclusively on promotional texts, other aspects of RC operation were not considered in this study. For instance, in the Netherlands, the Social Support Act (Wet maatschappelijke ondersteuning, Wmo) places responsibility on municipalities to help residents live independently and participate in society, creating incentives for them to fund RCs. Finally, cultural values are not static or monolithic (47); intra-country variation, including regional, institutional, or socio-economic differences, were beyond the scope of this analysis.

## Conclusion

This study identified how Recovery Colleges in the Netherlands characterise recovery in public-facing texts and how these narratives compare with those in England and Japan. Our findings indicate that Dutch RCs frame recovery as a socially embedded, everyday process of regaining agency, consistent with the country’s Quality-of-Life Orientation. In contrast to the individualised, education-driven messaging found in England, and the collectivist, meaning-making emphasis observed in Japan, Dutch RCs foreground relationality, self-direction, and societal contribution.

These culturally situated differences highlight how public representations of mental health recovery are shaped by broader societal values. By applying corpus-based methods to cross-national RC texts, this study contributes a novel linguistic and cultural lens to the understanding of recovery-oriented practice. Future research should examine how these textual framings align with lived experience and operational realities in different cultural contexts, supporting culturally responsive recovery models.

## Data Availability

All data underlying this study are openly available on the Open Science Framework at https://osf.io/9f7pv/.

All authors declare no conflict of interest.

All procedures followed were in accordance with the ethical standards of the responsible committee on human experimentation (institutional and national) and with the Helsinki Declaration of 1975, as revised in 2000 (5). Informed consent was obtained from all patients for being included in the study.

## Ethics statements

Patient consent for publication: Not applicable. Ethics approval: Not applicable.

## Acknowledgements

We would like to thank Nigel Henderson who helped facilitate the completion of RC surveys in Scotland. MS acknowledges the support of NIHR Nottingham Biomedical Research Centre.

## Supporting information

**S1 Supporting Information**. Methodological terms.

